# Reconstructing dengue transmission and assessing future outbreak risk in America Samoa, 2008–2023

**DOI:** 10.64898/2025.12.16.25342376

**Authors:** Sandra J. Kiplagat, Forrest K. Jones, Matt D.T. Hitchings, Scott Anesi, Angelynn Papu, Hans Desale, Janice Perez-Padilla, Emma S. Jones, Mark Delorey, Laura E. Adams, Gabriela Paz-Bailey, Joshua M. Wong, Adam Konrote

## Abstract

American Samoa experienced multiple dengue outbreaks during 2008–2023. We employed a catalytic model to reconstruct historical dengue virus (DENV) transmission dynamics by estimating the annual force of infection (FOI) – or probability of DENV infection among susceptible individuals – during this period. We incorporated data from 661 dengue cases between 2016–2023 and two serosurveys. We also modeled the proportion of the population with 0, 1, or ≥2 previous infections to assess population risk.

We estimated elevated FOIs during 2009 (11%; 95% CrI: 2%–38%), 2010 (15%; 95% CrI: 5%–29%), 2017 (50%; 95% CrI: 37%–67%), and 2018 (13%; 95% CrI 7%–24%). The proportion of the population with 0 previous infections was 22% in 2016, declined to 11% following the 2017–2018 outbreak, and rebounded to 20% in 2023. Our findings indicate a pattern of episodic dengue transmission in American Samoa, with substantial fluctuations in population immunity over time.

## Introduction

Dengue has emerged as a growing public health concern in the Western Pacific, with dengue outbreaks reported from multiple countries in 2025 (1). However, historical transmission patterns vary widely, with some areas experiencing large but sporadic outbreaks with no transmission detected in inter-epidemic periods, while others have established endemic transmission or co-circulation of multiple serotypes (2, 3). Dengue is caused by four antigenically distinct but closely related dengue viruses (DENV-1, DENV-2, DENV-3 and DENV-4), also called serotypes (4). DENV infections are often asymptomatic or can cause mild, non-specific illness; however, dengue can cause severe and life-threatening disease. The risk for severe disease is influenced by many factors, including age and coexisting conditions such as hypertension, diabetes, asthma, or pregnancy. Immune status and previous exposure to DENV are also to play a role. Individuals without prior DENV infection (DENV-naïve) have a moderate risk for severe dengue during the first (primary) infection. Individuals who become infected with a different serotype after a prior infection (a secondary infection) have a higher risk for severe dengue due to antibody dependent enhancement (5). Although severe dengue is also possible during post-secondary infections (third or fourth infections), the risk is lower during this period as people have developed a more protective immune response (5).

American Samoa, an unincorporated territory of the United States, is characterized by large but sporadic dengue outbreaks (3), distinct from the more persistent endemic transmission patterns observed in many countries in Latin America and Asia (6). However, DENV transmission in American Samoa still meets the Yellow Book criteria for frequent or continuous transmission, defined as reporting more than 10 cases in at least three of the previous ten years (7). The earliest reported dengue outbreak in American Samoa occurred in 1911 (8). In the past two decades, dengue outbreaks occurred in 2009–2010 with >400 cases (9) (predominantly DENV-4) and 2015 with 898 cases (predominantly DENV-3) (10). A large outbreak during 2017–2018 included 1,081 laboratory-confirmed cases (predominantly DENV-2) (11). During 2019–2024, American Samoa reported only one locally acquired case. On July 8 2025, the American Samoa Department of Health declared an outbreak after two locally acquired dengue cases were detected. As of December 1, over 300 dengue cases have been reported in American Samoa in 2025 (1).

Dengue case surveillance is valuable to describe disease burden and transmission dynamics over geography and time. However, traditional surveillance underestimates true transmission rates because asymptomatic infections, which comprise nearly three-quarters of all DENV infections, go undetected (12). Additionally, not all symptomatic cases seek care, and appropriate testing is not always performed or available (13, 14). In American Samoa, it is unclear whether the years with no dengue cases reported reflect a true absence of DENV transmission or under-reporting through passive surveillance systems during non-epidemic periods.

Serological surveillance can provide a clearer picture of historical DENV transmission (15) by revealing the cumulative exposure to DENV, including asymptomatic or subclinical infections missed by case surveillance (12, 16).This can be achieved by estimating the annual force of infection (FOI), defined as the probability of infection with any DENV serotype among DENV-naïve individuals within a year. Catalytic models, when applied to age-stratified serological data, can estimate FOI over time, even with limited surveillance data. Additionally, incorporating individual residency histories can help identify variations in exposure risk, providing more precise estimates of population immunity and transmission intensity (17, 18). Annual FOI estimates are also useful for modeling population immunity (e.g., the proportion of population who are DENV-naïve, have experienced only one previous DENV infection, or who have experienced multiple DENV infections). This can inform understanding of the potential for future outbreaks and design intervention programs (e.g., vaccination) to protect age groups at greatest risk.

We developed an integrated analytic approach that combines dengue case data and age-stratified seroprevalence surveys within a catalytic modeling framework to reconstruct dengue transmission patterns from 2008 to 2023. This analysis aims to: 1) determine if there is evidence of local transmission of dengue in American Samoa when no cases were officially reported 2) compare current population immunity to dengue immunity before the previous outbreak, and 3) identify the age groups currently at highest risk for severe dengue in American Samoa.

## Methods

### Data Sources

#### Case-based Surveillance Data

U.S. states and territories report dengue cases to CDC through the National Arboviral Surveillance System (ArboNET), which collects detailed information on each case, including demographics, dengue serotype, clinical characteristics, case classification, and whether a case was travel-associated or locally acquired. Dengue became a nationally notifiable disease in 2010. The dengue case definition used in this analysis was established by the Council of State and Territorial Epidemiologists in 2015, and included clinical criteria (fever and one of the following symptoms: nausea/vomiting, rash, aches and pains, tourniquet test positive, leukopenia), epidemiologic criteria (e.g., residence in or travel to an area with ongoing dengue transmission within the preceding two weeks of onset of an acute febrile illness), and confirmed or probable laboratory evidence of DENV infection. For this analysis, we included dengue cases in American Samoa that met the case definition and were reported to ArboNET from 2016 through 2023. Although 1,081 laboratory-confirmed cases were previously reported (11), only 661 cases were captured in ArboNET (19) and therefore included in the analysis.

#### Serosurvey Data

We used data from two dengue serological surveys in American Samoa for this analysis. In 2010, Duncombe and colleagues conducted a household-based dengue seroprevalence study in American Samoa, testing 794 adults aged 18–87 years for dengue IgG antibodies (20). Participants were recruited from Tutuila (where >97% of American Samoa’s population lives), Aunu’u, and Manu’a islands. On Tutuila and Aunu’u, spatial sampling selected one adult per household; in Manu’a islands, convenience sampling approach was used due to smaller village sizes. For our analysis of transmission patterns, we included data from a serosurvey conducted only among individuals aged 18–40 years old from all islands (n=418).

The second serologic survey (21) was a school-based cross-sectional dengue serosurvey among 887 children and adolescents in Tutuila, American Samoa during September–October 2023 (21). Seven of 36 public schools were randomly selected through a single-stage cluster sampling design, stratified by school type (elementary and junior high school). All students in grades 3–10 (ages 7–16) from selected schools were invited to participate. Information was collected on demographics, birthplace and residency information throughout their lifetime. Students were tested using the CTK Biotech OnSite dengue immunoglobulin G (IgG) rapid test (sensitivity = 89.6%; specificity = 95.7%) (22). For this analysis, we removed 24 participants with indeterminate serological results out of 887 tested.

#### Historical Residency Data from 2023 Serosurvey

Using the responses from the survey questionnaire, we reconstructed historical residency profiles for each individual by incorporating country of birth, country of residence, and years and locations lived outside of American Samoa. Specifically, we identified the location where each individual lived for every year of their life (see Supplementary Methods). For each year of life for each person, we classified the residency locations outside of American Samoa according to the CDC Yellow Book dengue classifications (7), where locations with “frequent or continuous” risk were classified as endemic, while areas with “sporadic or uncertain” or “no evidence” of risk were classified as non-endemic.

### Statistical Analysis

Estimates of seroprevalence and 95% confidence intervals were computed using the survey package (23) in R version 4.4.1 (24). All point and interval estimates incorporate design weights reflecting the school enrollment and proportion of eligible grade levels to account for the complex sampling design and ensure population-level representativeness. Estimates and intervals were adjusted for imperfect testing, assuming a sensitivity of 89.6% and specificity of 95.7% (22). A quasi-binomial generalized linear model was used to evaluate statistical significance of the difference between the location categories. Pearson’s chi-squared asymptotic exact test was used to compare the proportion of seropositive individuals born in non-endemic areas that arrived after the outbreak to that of seronegative individuals born in non-endemic areas.

We developed a catalytic model framework integrating serological survey data, age-stratified dengue case data, and individual residence histories to estimate the annual dengue FOI in American Samoa (Supplementary Methods) as well as the average FOI among people previously residing outside American Samoa at any time, regardless of the duration of their residence, whether in endemic and non-endemic areas. Briefly, the model extends methods by Kada et al. (25), incorporating random effects for annual variation in reporting rates and school-specific risk. We modeled seropositivity as a function of cumulative infection hazard and adjusted for test sensitivity and specificity. We allowed cases to be a mix of both primary and secondary infections which each had different reporting rates.

We fit the model using Stan and implemented with the rstan package in R. We ran 4 independent Markov Chain Monte Carlo (MCMC) chains, each with 1000 iterations (500 burn-in and 500 sampling). We assessed convergence using visual inspections of traceplots and the potential scale reduction factor (R-hat) (26). We conducted posterior predictive checks to ensure the model captured the observed age-specific patterns with case-based surveillance and seroprevalence, supporting the model’s reliability in estimating FOI. We reported the results as median estimates and 95% credible intervals (CrI). We report the FOI as an annual hazard probability (i.e., the probability of infection with any dengue serotype given susceptibility). Susceptibility is determined by an individual’s infection history: DENV-naïve individuals are susceptible to all four serotypes; those with a single prior DENV infection have lifelong immunity to that specific serotype but remain susceptible to the others; and individuals with multiple prior infections have acquired broader immunity. We also extracted MCMC draws to estimate the average annual FOI in American Samoa, the proportion of the population with different DENV infection histories (i.e., DENV-naïve, one previous infection, or multiple DENV infections) at the end of each year (and their ratios), and the proportion of individuals in five-year age groups that were infected only once with DENV at the end of 2023.

Dengue risk could vary by residency outside American Samoa. We examined the sensitivity of the annual FOI estimates to assumptions of the reported residency for individuals in the 2023 serosurvey. First, we examined how our choice of residency for individuals who reported in two locations within the same year changed our FOI estimates. This is important because individuals with dual residency may have varying exposure to dengue transmission dynamics in each location, which could influence their overall risk of infection, and subsequently the FOI estimates. We did this by comparing FOI estimates calculated using the first (i.e., former) reported location and second (i.e., latter) reported location for these individuals. We also compared these FOI estimates to those calculated with the assumption that all individuals lived in American Samoa their entire lifetime (regardless of their true residency history) and restricting the analysis to individuals who had resided in American Samoa throughout their lifetime.

Secondly, we interrogated our model’s assumption that each year a mixture of serotypes circulated; in reality, the 2017–2018 epidemic was dominated by DENV-2 and the susceptibility profile of the population in 2018 was likely lower than estimated. To assess the sensitivity of our estimates to this assumption, we collapsed year 2017 and 2018 into a single epidemic year.

Code presented in this study is available at https://github.com/fjones2222/dengue-americansamoa-catalytic.

## Results

### Revisiting results from dengue surveillance data in American Samoa

During 2016–2023, 661 cases were reported to ArboNET, 658 (99.5%) of which occurred during 2017–2018 (Fig. 1). Most (60.2%) were under 18 years of age and half (50.2%) of the cases were female. Most cases (58.2%) were reported from the Western district of American Samoa. Most cases had an unknown serotype (81.4%), and only DENV-2 was detected in 122 samples with known serotype. Nearly half of cases (45.4%) were hospitalized for DENV infection. No fatalities were reported. Nearly all cases were classified as locally acquired (99%), with only 1 travel-associated case in 2016 and 1 locally acquired case in 2022.

**Figure 1:**
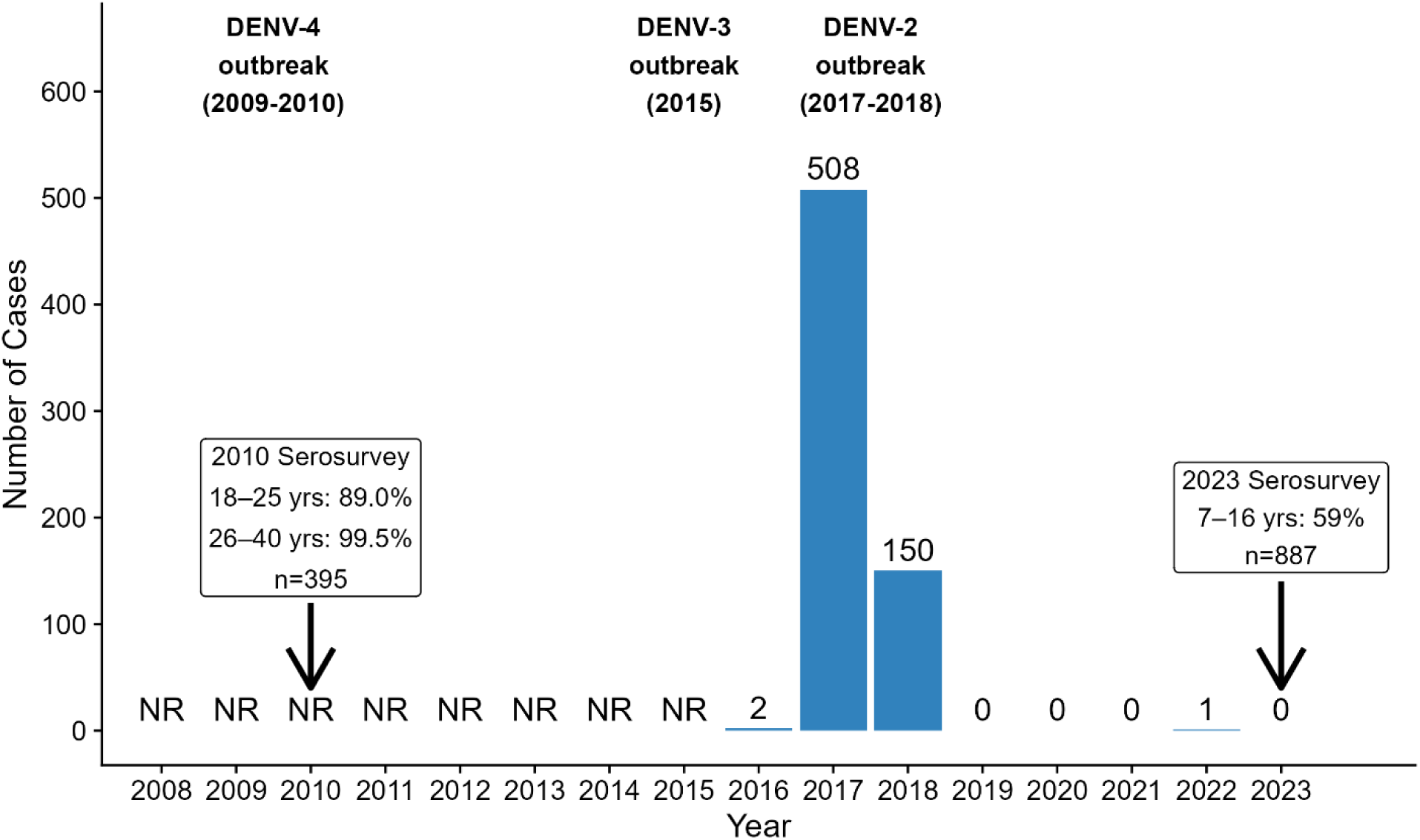
Available dengue case data and serosurvey data, American Samoa, 2008-2023. An epidemic curve of cases reported to ArboNET between 2016 and 2023 is shown with blue bars. NR = Cases were not reported to ArboNET before 2016. Timing and results of IgG serosurveys are shown in boxes with arrows. Timing and serotype information of recent dengue outbreaks identified from published literature are shown in bold.

In the 2010 household serosurvey (20), a total of 418 adults aged 18–40 years were tested for previous dengue virus infection. Among the participants, 89.1% aged 18–25 and 99.5% aged 26–40 years tested positive for previous dengue virus infection.

In the 2023 school-based serosurvey, a total of 887 (39%) students out of 2,267 invited students participated. The estimated seroprevalence was 59% (95% CI: 47–70) for all students, as previously reported (21). After we excluded individuals with uninterpretable results (n=24) or missing residency information (n=3), 860 participants remained for the analysis (Fig. S1). The median age was 11 years (range: 7–16 years) and more than half (54%) were female (Table S1). Of the total tests remaining, 490 (57%) were seropositive and 370 (43%) were seronegative.

In the 2023 serosurvey, 746 (87%) were born in American Samoa, 76 (9%) in endemic areas (including Samoa, Fiji, Philippines, and Tuvalu) and 38 (4%) in non-endemic areas (continental United States, Hawaii, Australia, and New Zealand) (Table S1). There were no significant differences in seroprevalence by location of birth (Fig. S2), although seropositive children born in non-endemic areas were more likely to have moved to American Samoa prior to or during the 2017–2018 outbreak than seronegative children born in non-endemic areas (95% vs 61%, p=0.004, Fig. S3).

### Dynamics and immune landscape of dengue in American Samoa from the catalytic model

After fitting the catalytic model, we observed a reasonable fit to the serosurvey results of 2023 (Fig. 2A) and the 2016–2023 case surveillance data, both in terms of the number of cases per year (Fig. 2B) as well as the overall age distribution (Fig. 2C). We estimated the variance of the multiplicative random effect (i.e., 1/α as described in the Supplementary Methods) for school of serosurvey participant to be 0.42 (95% CrI: 0.34–0.88).

**Figure 2:**
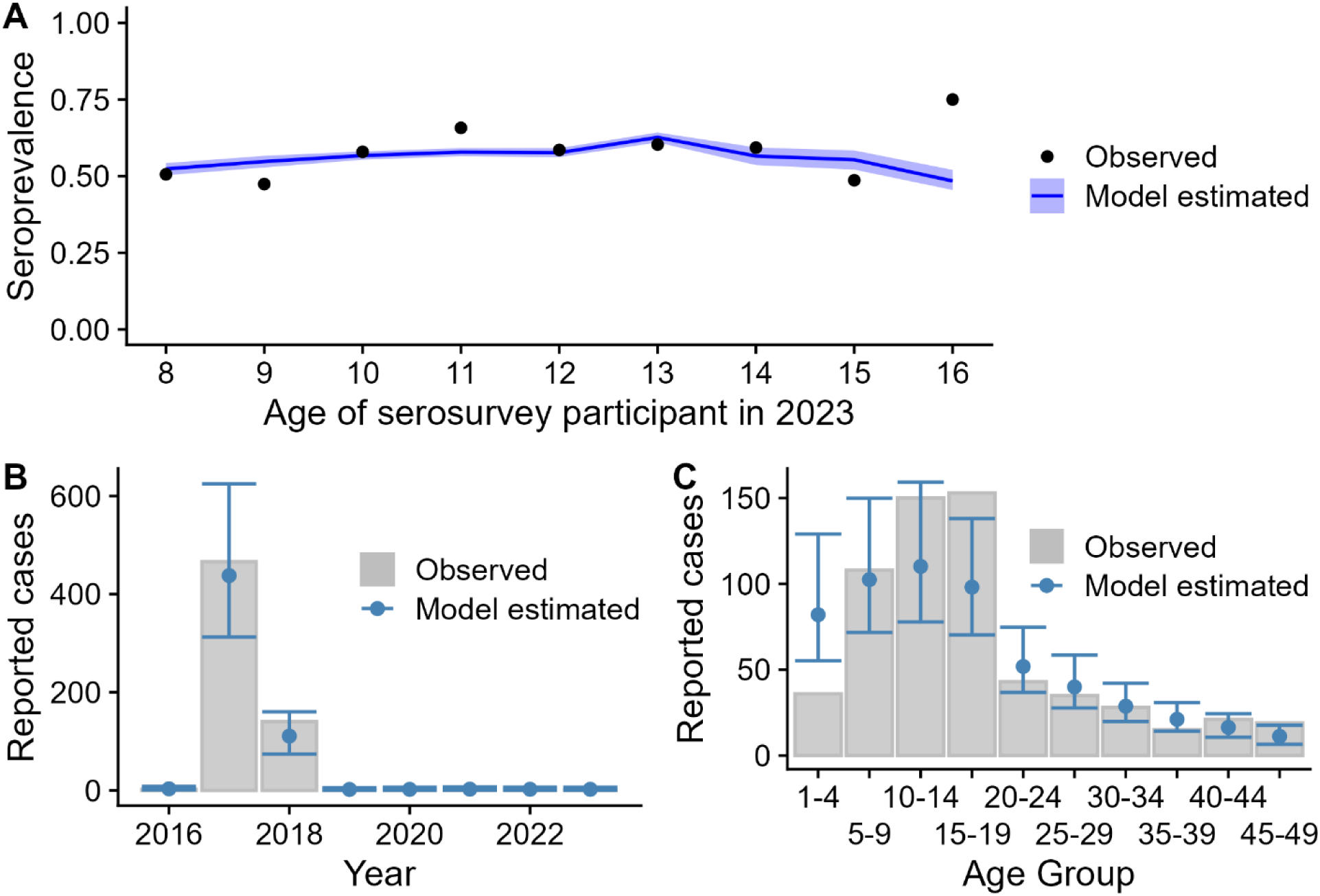
Assessment of catalytic model fit to serological and case data. (A) Temporal trends in model-estimated dengue seroprevalence in 2023 (blue line and blue shading) compared to observed seroprevalence data points from serosurvey conducted in 2023. As dengue outbreaks are sporadic and the last dengue outbreak was in 2017, children have had similar exposure histories, resulting in similar seroprevalence by age. (B) Annual reported dengue cases from American Samoa surveillance data submitted to CDC ArboNET, 2016–2023. Gray bars represent the number of cases reported to ArboNET. Blue points represent the model estimated number of cases per year, and blue lines represent the 95% Credible Interval (CrI). (C) Age distribution of reported dengue cases from surveillance data submitted to CDC ArboNET during 2016–2023. Gray bars represent cases reported to ArboNET. Blue points represent the model estimated number of reported cases per age group, and blue lines represent the 95% CrI.

Elevated FOI were observed in 2009 (11%; 95% CrI: 2%–38%) and 2010 (15%; 95% CrI: 5%–29%) when a previous outbreak had been reported in an epidemiological update but not reported in ArboNET (9, 27). Between 2011 and 2016, the annual FOI was low, ranging from 2% to 8%. The annual FOI was highest during 2017 (50%; 95% CrI: 37% –67%) and 2018 (13%; 95% CrI 7%–24%), aligning with reported high case numbers during the outbreak (Fig. 3A). Between 2019 and 2023, every year the FOI was estimated to be (1%), which was not statistically significantly different from model-estimated FOI in non-endemic areas (Table S2). This suggests that there was little to no DENV transmission between 2019 and 2023. We also estimated the average annual FOI during 2008–2023 to be 9% (95% CrI: 7%–14%) for American Samoa, 0.6% (95% CrI: 0.3%–2%) for people residing in non-endemic areas, and 15% (95% CrI: 10%–24%) (Fig. 3B) for other people residing in endemic areas.

**Figure 3:**
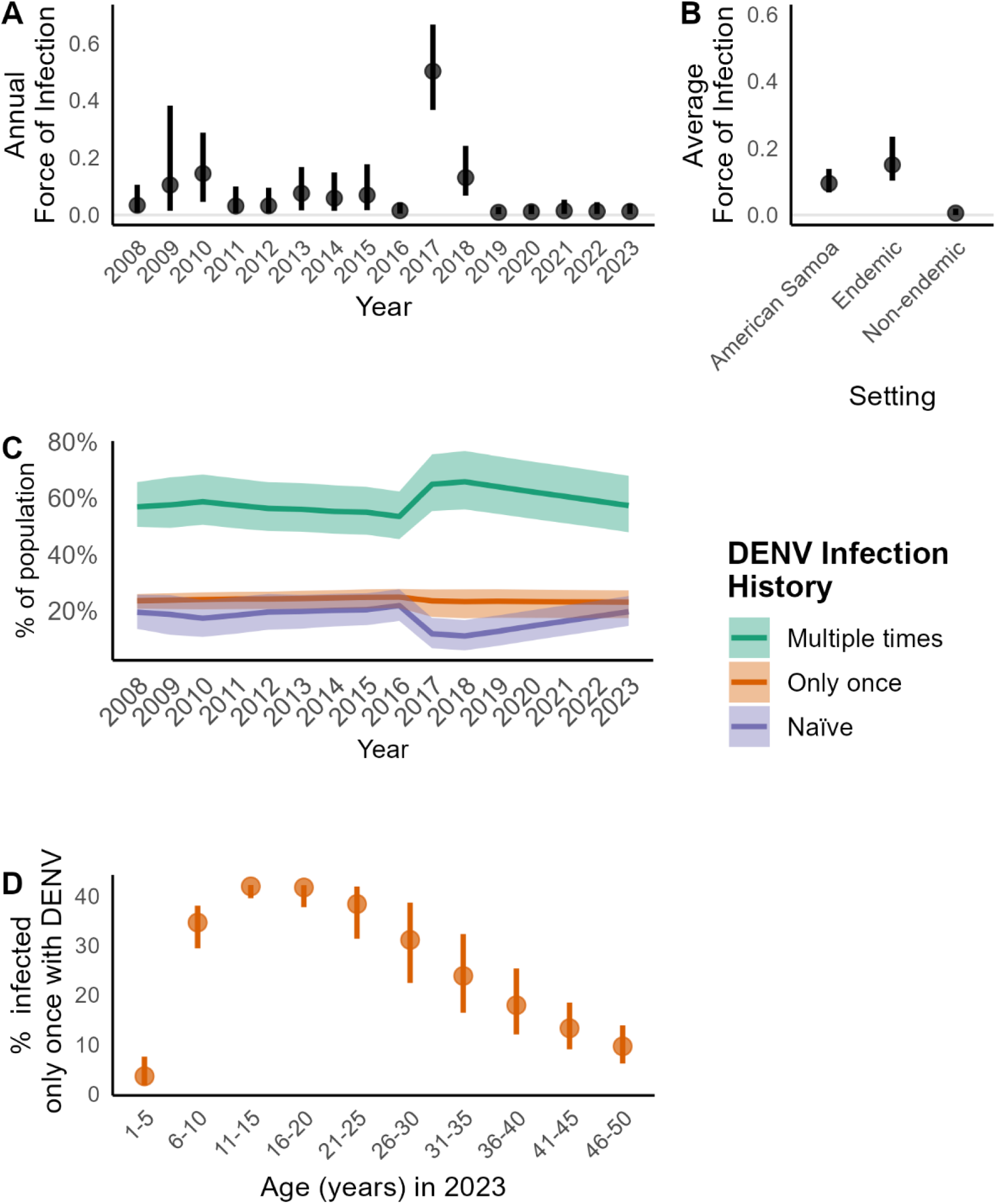
Reconstruction of dengue transmission and risk from catalytic model. (A) Estimates of the annual force of infection in American Samoa (black dots) with 95% Credible Intervals (black lines). (B) Estimates of the average annual force of infection in American Samoa, endemic areas, and non-endemic areas history, 2008–2023. The light grey horizontal line shows the force of infection of zero. (C) Estimated DENV infection history in the American Samoan population by year, with proportion of people who had not experienced a DENV infection (DENV-naïve) shown in purple lines and shading, the proportion of people with a single DENV infection (primary infections) in orange lines and shading, and the proportion of people with multiple (>1) previous DENV infections in green lines and shading (D) Estimated proportion of the population with one previous DENV infection (orange dots) by age in 5 year intervals, indicating the group that would be at higher risk for severe disease on a second infection.

Our model identified a significant shift in the dengue immunity landscape in American Samoa after the 2017–2018 outbreak. At the end of 2016 (i.e. before the 2017–2018 outbreak), an estimated 22% (95% CrI 16%–28%) of the population had never been infected with DENV (DENV-naïve), 25% (95% CrI 21%–28%) had been infected once with DENV, and 54% (95% CrI 45%– 62%) of the population had been infected with two or more DENVs (Fig. 3C). At the end of 2018, the proportion of individuals who were DENV-naïve had decreased to 11%, which was 0.51 (95% CrI: 0.33–0.67) times that in 2016. The proportion of DENV-naïve people in the population gradually increased during the low transmission years of 2019–2023, reaching 20% (95% CrI: 15%–25%) in 2023, similar to the levels in 2016 before the 2017–2018 outbreak. The proportion of individuals with a primary DENV infection remained stable from 2008 to 2023, ranging from 23% to 25%. The proportion of the population with multiple previous DENV infections ranged from 54%–59% during 2008–2016, increased during the outbreak to 66%, and declined to 57% from 2019–2023.

To identify age groups that may be at higher risk for severe disease, we used the catalytic model to estimate the proportion of the population with one previous DENV infection at the end of the analysis period (2023). The highest proportion of primary infections occurred among adolescents aged 11–20 years old (42%). The proportion of the population with one previous DENV infection decreased after age 20, with 10%–23% of adults aged 30–50 years having one previous DENV infection, as a higher proportion had been infected with DENV multiple times. The age group with the lowest DENV exposure occurred among children younger than 5 years old, with an estimated 4% (95% CrI: 2%–8%) having experienced a single previous DENV infection.

We conducted sensitivity analyses to understand how much residency data impacted our findings. Overall, we found limited variation in the annual estimated FOI for most years by residency data used (Fig. S4), although limiting the dataset only to those with lifelong residency resulted in an increase of the uncertainty in the 2016 FOI estimate. Findings were similar to the annual estimated FOI and the proportion of individuals in each infection history category if we assumed 2017 and 2018 was a single epidemic year.

## Discussion

Our findings suggest DENV transmission in American Samoa is characterized by episodic, outbreak-driven dynamics rather than sustained endemic transmission. We estimated the highest FOI during the 2017–2018 outbreak, consistent with high case numbers and incidence reported through case surveillance. During 2019–2023, the estimated FOI was similar to non-endemic areas, suggesting that local DENV transmission may have stopped (or only occurred at very low levels) after the outbreak ended in 2018. The model also estimated elevated FOIs aligning with the outbreak in 2009–2010, with very low inter-epidemic transmission, supporting the hypothesis that periodic viral re-introductions result in population-wide outbreaks (9–11). This pattern is consistent with dengue epidemiology in other Pacific Island nations, where outbreaks occur every 3–5 years due to virus introductions, ecological shifts, or waning immunity (2, 3). The regional consistency indicates the island geography, vector abundance, and travel patterns may support conditions that favor episodic compared to endemic transmission, in contrast to larger geographic areas where viral circulation is maintained at low levels in an endemic cycle (9, 28, 29).

The 2017–2018 outbreak reshaped the dengue immunity landscape in American Samoa. Our model estimated that the proportion of individuals who had never been exposed to dengue was nearly cut in half after the 2017–2018 outbreak, consistent with the FOI of 50%. However, in 2023 (after several years without documented local transmission), our model suggests that population-level immunity had returned to pre-outbreak levels, increasing the risk for a new dengue outbreak in America Samoa. American Samoa’s dengue outbreak in 2025 may be occurring due to several factors, such as frequent DENV introductions from other Pacific Island nations (such as Samoa) experiencing outbreaks in 2025 (1).

We also found that individuals 11–20 years old (in 2023) had the highest prevalence of only one DENV infection in their lifetime. These individuals stand to benefit the most from dengue vaccination to prevent hospitalization and severe dengue from a subsequent DENV infection. The episodic transmission pattern observed in American Samoa means that seroprevalence by age can change over time depending on the time between outbreaks. Health departments in locations with similar episodic transmission patterns should consider these dynamics when designing potential vaccination programs. As seroprevalence by age has been an important factor in the recommendations for both dengue vaccines evaluated by the World Health Organization to date (Dengvaxia and Qdenga) (30), updated seroprevalence estimates may be needed in American Samoa and other areas with episodic transmission when making decisions around vaccine implementation.

Our analysis of residency histories from the 2023 serological survey (particularly from those spending ≥1 year in non-endemic areas history) provided valuable insights into local transmission in American Samoa. We found that the average FOI in American Samoa was significantly higher than in non-endemic areas, but significantly lower than in endemic areas (principally Samoa as more students spent time there than other areas outside of American Samoa). Samoa reported dengue cases in 2022, 2023, and 2024, in contrast to American Samoa which reported only a single case to ArboNET for those years (31). The differences in the FOI estimates for American Samoa and Samoa suggest that there was a lower risk of DENV infection in American Samoa, not just a potential difference in sensitivity of the surveillance systems. We also found that including residency data in the catalytic model did not dramatically change our annual estimates of the FOI, likely because most students (84%) spent their entire lives in American Samoa. However, by systematically collecting this comprehensive data, we were able to better understand the nuances and complexity of dengue exposure throughout an individual’s life and improve estimates of dengue transmission in American Samoa.

This analysis has some limitations. FOI estimates from 2008–2015 should be interpreted with caution as no case surveillance data were available in ArboNET during this period, despite the occurrence of two dengue outbreaks. Given the missing case data, we were not surprised the model did not estimate a large FOI in 2015 and may have distributed that risk across the years 2013 to 2015 (which were estimated to have moderate FOI levels). Although we found data on residency history to be useful for this study, we had to make imperfect assumptions based on the data collection procedure (see Supplementary Methods).While the model captures residency-based seroprevalence patterns that align with known outbreak years, these data do not include information about short term travel to areas where a participant could have been infected with dengue. Additionally, our analysis did not attempt to reconstruct the transmission dynamics of individual serotypes. Future serosurveys in American Samoa might consider measuring serotype-specific antibodies to further refine the reconstruction of dengue dynamics. Lastly, for estimates of seroprevalence from the 2023 serosurvey, calibration and post-stratification were not conducted, as the sampled population closely aligned with the population distributions on available demographic variables such that adjustment beyond accounting for sampling weights was deemed unnecessary.

Our study’s approach to integrate case surveillance data and serological data across multiple years can be valuable in settings that have limited surveillance data or where dengue outbreaks are infrequent. Quantifying the dynamics of population immunity in settings with episodic dengue transmission (like American Samoa) has implications for understanding risk for future outbreaks and age groups at highest risk for secondary infection. These findings can inform preparedness plans and direct prevention efforts (such as vaccination programs) to age groups at heightened risk for severe disease.

## Data Availability

All data produced in the present study are available upon reasonable request to the authors

## Acknowledgments

We thank Astrid M. Johannson, Noelle Tavale, Annette F. Ilimaleota, Jacki M. Tulafono, and Motusa Tuileama Nua (ASDOH) as well as Emi Chutaro and Janet Camacho (Pacific Island Hawaii Office Association) for their contributions to implementing the serological survey in 2023. We also thank Tyler Sharp for providing background knowledge from during the dengue outbreak investigation in American Samoa in 2015.

## Supplementary Tables

**Supplementary Table 1.**
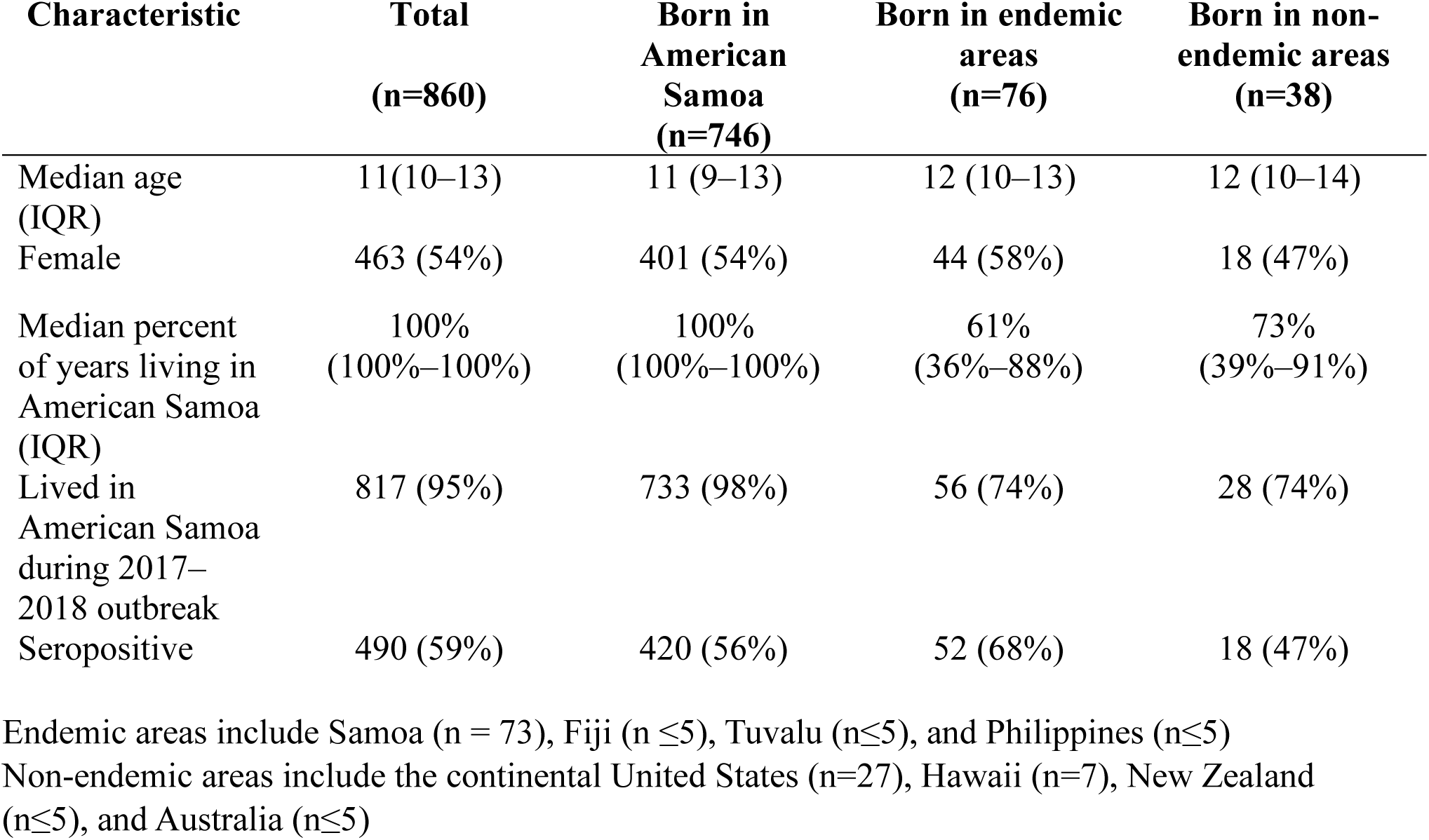
Demographic characteristics among participants in a dengue serosurvey in American Samoa by birth location, September-October 2023.

| **Characteristic** | **Total**<br>**(n=860)** | **Born in**<br>**American**<br>**Samoa**<br>**(n=746)** | **Born in endemic**<br>**areas**<br>**(n=76)** | **Born in non-**<br>**endemic areas**<br>**(n=38)** |
| --- | --- | --- | --- | --- |
| Median age<br>(IQR) | 11(10–13) | 11 (9–13) | 12 (10–13) | 12 (10–14) |
| Female | 463 (54%) | 401 (54%) | 44 (58%) | 18 (47%) |
| Median percent<br>of years living in<br>American Samoa<br>(IQR) | 100%<br>(100%–100%) | 100%<br>(100%–100%) | 61%<br>(36%–88%) | 73%<br>(39%–91%) |
| Lived in<br>American Samoa<br>during 2017–<br>2018 outbreak | 817 (95%) | 733 (98%) | 56 (74%) | 28 (74%) |
| Seropositive | 490 (59%) | 420 (56%) | 52 (68%) | 18 (47%) |

Endemic areas include Samoa (n = 73), Fiji (n ≤5), Tuvalu (n≤5), and Philippines (n≤5)
Non-endemic areas include the continental United States (n=27), Hawaii (n=7), New Zealand (n≤5), and Australia (n≤5)

**Supplementary Table 2.** Variables Used for Residency Data Cleaning.

| Variable Name | Survey Label | Purpose / Use | Notes |
| --- | --- | --- | --- |
| Date of birth | Date of Birth | Determines year of birth and timeline start | Used to calculate year of birth |
| 4. Where was your child born? | Birthplace | Primary source for birthplace classification | Often listed as “American Samoa” |
| Where was your child born?<br>Other: | Birthplace<br>(Other) | Clarifies birthplace not covered in dropdown | Used to identify other country |
| 5. My child has been a resident of American Samoa | Lifetime AS Residency Indicator | Distinguishes lifelong vs. partial residents | “All his/her life” or “Since __ year __” |
| My child has been a resident of American Samoa since (year) | Year Residency Began | Establishes year of AS residency for non-lifelong residents | Often missing for lifelong residents |
| If your child has lived somewhere else... (multiple) | Locations before AS residency | Identifies locations prior to AS | Used in location matrix |
| Residence in [residence_other], year start/end | Time-specific location history | Captures movements across years | Basis for constructing matrices |

**Supplementary Table 3.**
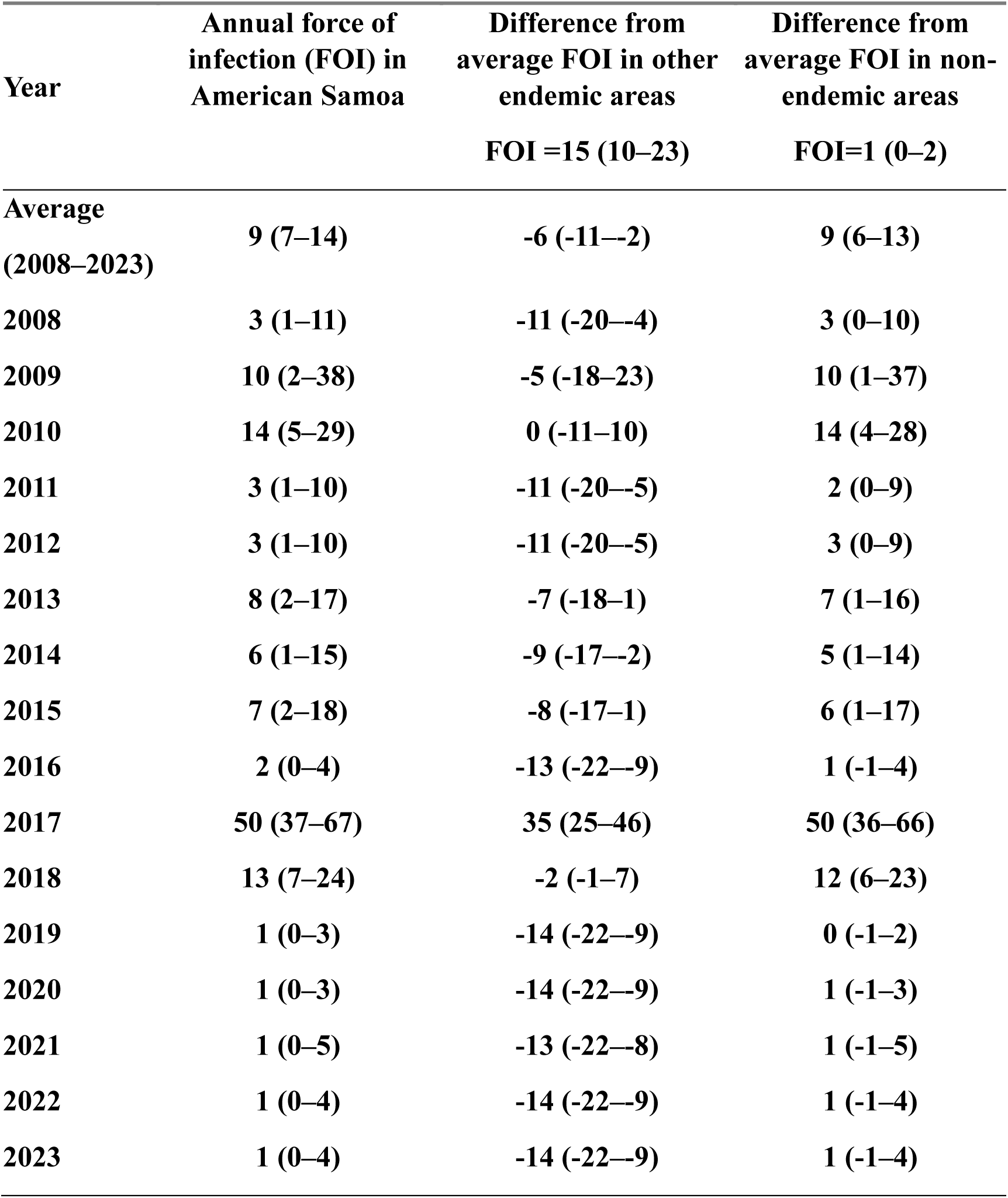
Differences (in percent) in the estimates in the force of infection by residency location.

## Supplementary Figures

**Supplementary Figure 1:**
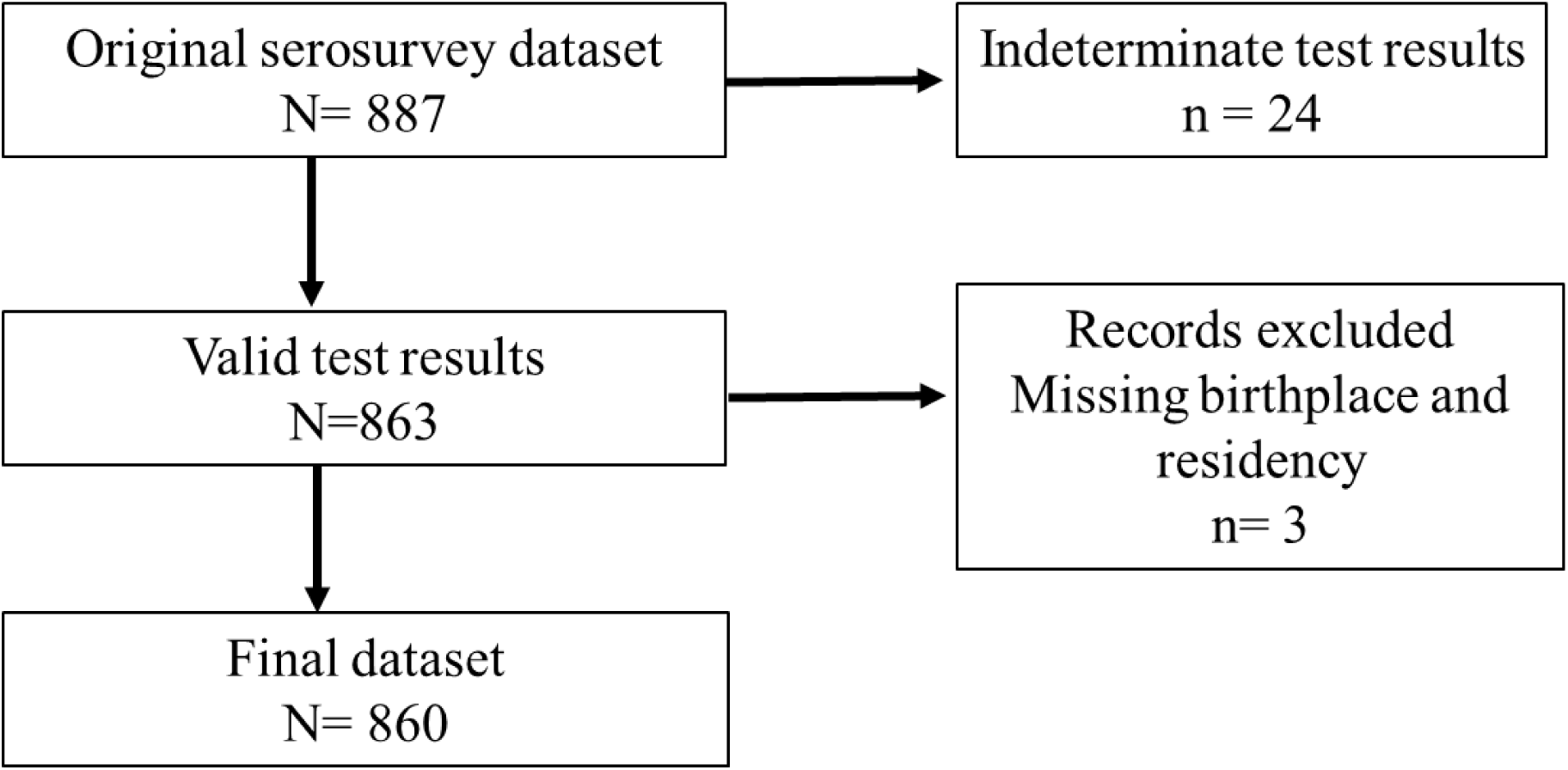
Participant selection from the school-based serosurvey, American Samoa, 2023.

**Supplementary Figure 2:**
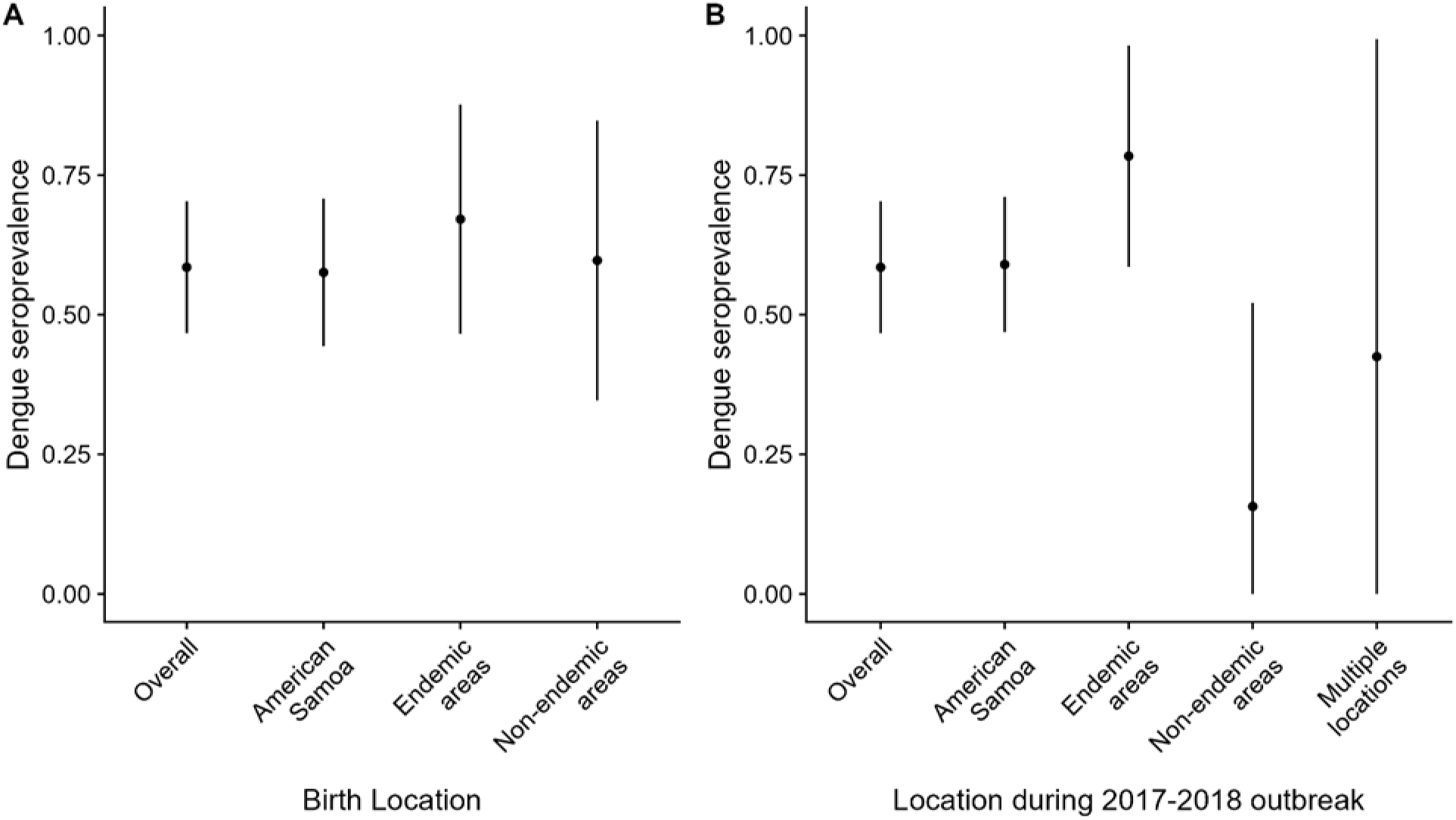
Modeled prevalence of past dengue virus infection among school children in 2023 serosurvey by place of birth and reported residency during the 2017 to 2018 outbreak—American Samoa, 2023. Seroprevalence estimates by (A) birth location and (B) reported residency location during the 2017–2018 outbreak. Multiple locations refer to individuals that had a different location of reported residency in 2017 and 2018.

**Supplementary Figure 3:**
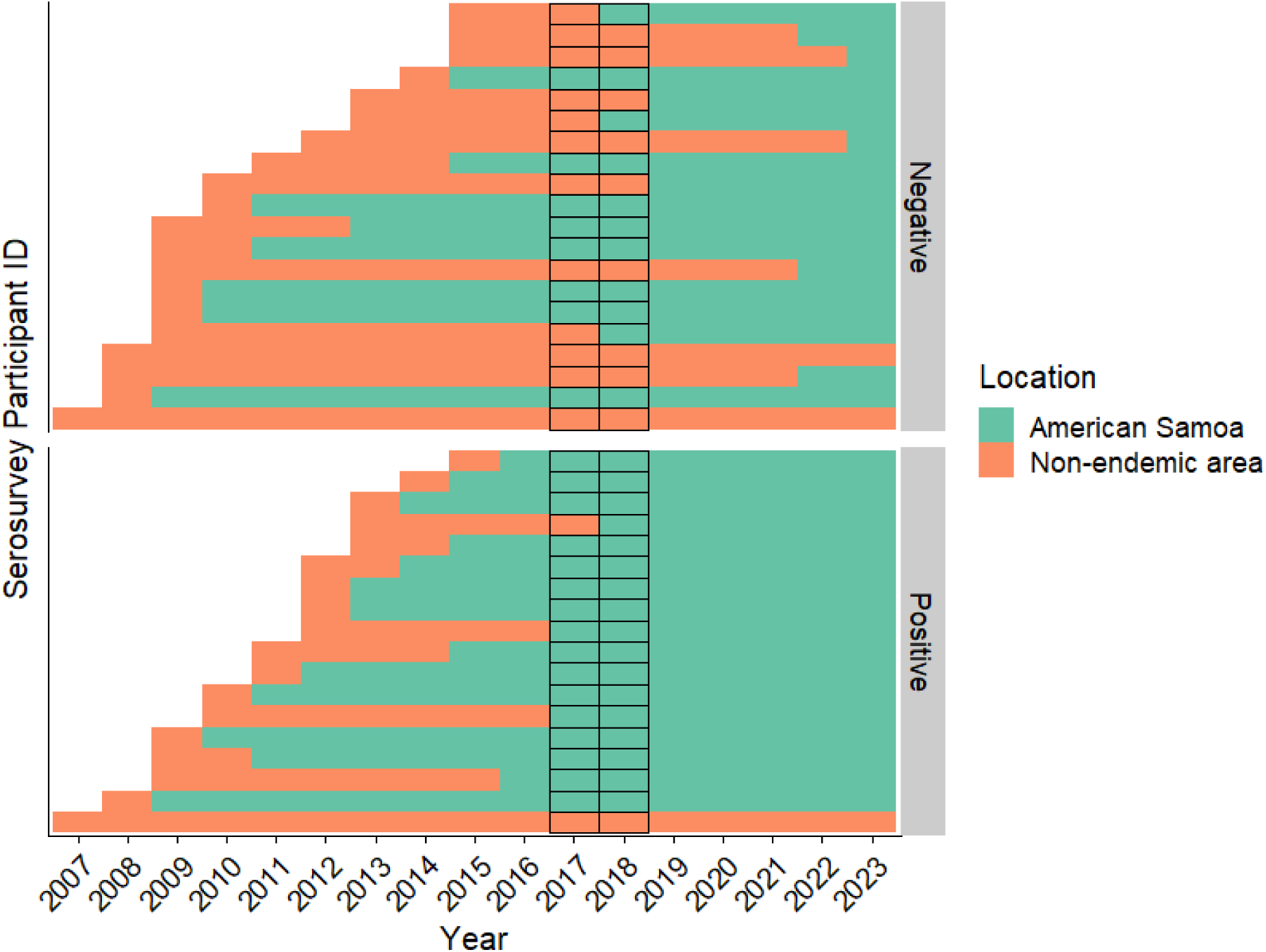
Residency timelines for individuals born in non-endemic areas history. Each row represents a participant who was born in an area with limited or no transmission history and each box represents the location of the participant on a particular year. The serostatus of the participants is shown in the grey box to the right. Black outlines indicate the outbreak period of 2017 and 2018.

**Supplementary Figure 4:**
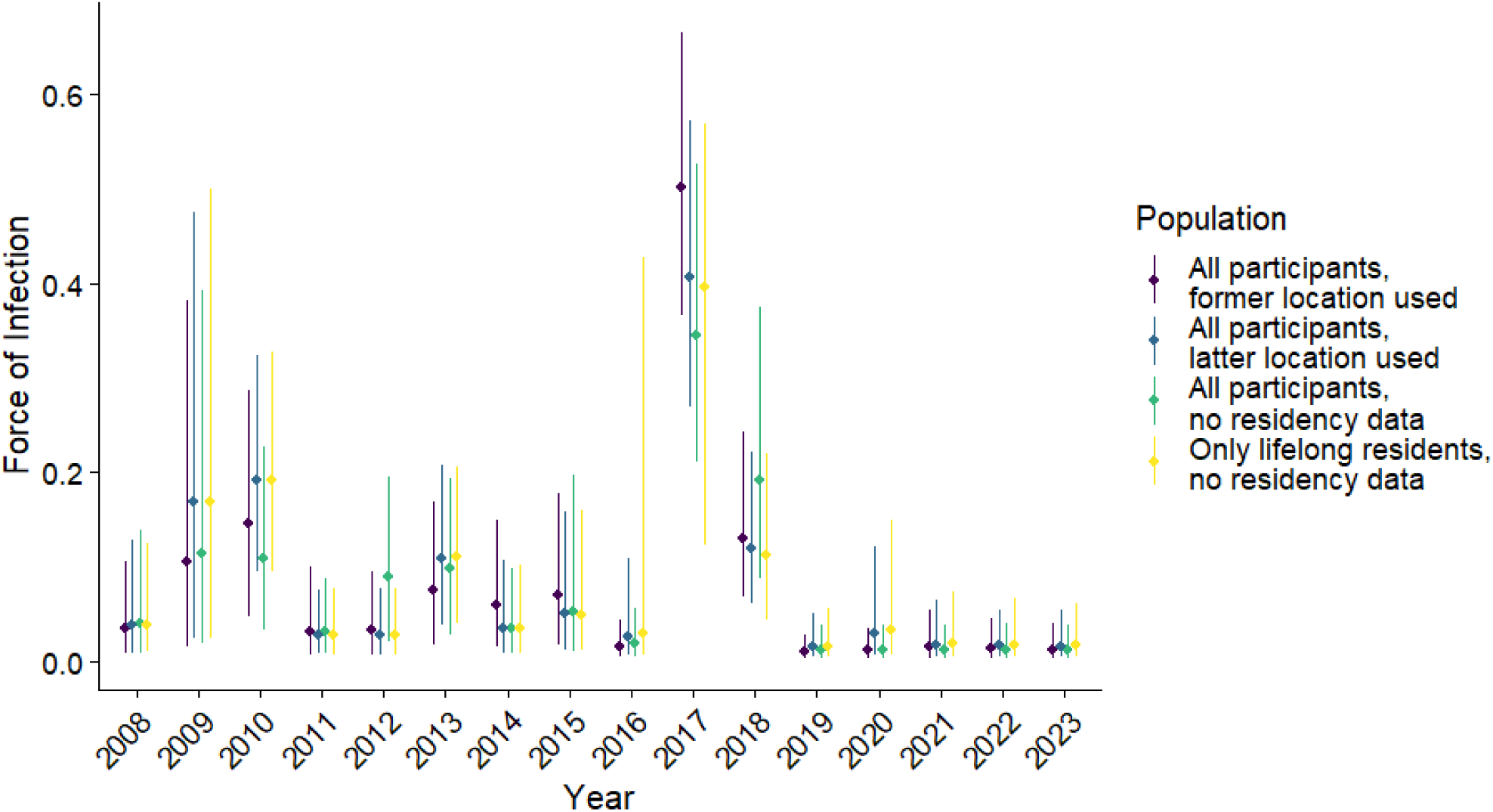
Sensitivity analysis of annual force of infections estimates using different assumptions of residency status. Annual FOI estimates are shown as point estimates with accompanying uncertainty intervals represented by lines. Four residency classification approaches were compared. Purple comprised of all participants assigning residency by former location for those who reported living in more than one location. Blue represents all participants, assigning residency by latter location for those who reported residing in more than one location. Green represents all participants without incorporating residency data. Yellow represents individuals who have lived in American Samoa throughout their life.

## Supplementary Methods

### Residency Data Cleaning

Initial data cleaning involved standardizing birthplace responses. Continental U.S. states (e.g. Tennessee, Alaska or Utah) were grouped under “Continental US”, while cities or islands in Hawaii were classified separately as “Hawaii.” Islands and villages within Pacific nations (e.g., “Upolu,” “Saipipi”) were grouped under their corresponding countries, such as “Samoa.”

### Residency Categories

Participants were classified into two major categories based on residency patterns:

- Lifelong American Samoa Residents: Individuals who were born in American Samoa and continuously lived there since birth.
- Partial American Samoa Residents: Individuals who either (a) moved to American Samoa after being born elsewhere, or (b) were born in American Samoa but had spent time living in a different location. For these individuals, the detailed location history was essential to reconstruct their residency profiles.

For individuals with lifelong American Samoa residency, their birthplace and year-by-year were listed as American Samoa. For partial residents, we constructed year-by-year residency from bith to 2023. Individuals with missing information on birthplace or residency were excluded from the analysis (n = 3; Fig. S1). The key variables used in cleaning and residency classification are summarized in Table S2. Residency timelines were constructed to capture annual changes in location and to enable exposure assessment based on place of residence. The classification allowed for identification of individuals with uninterrupted residency as well as with more complex residency profiles.

### Description of Former and Latter Datasets

To account for multiple residential histories within the same calendar year, we constructed year-by-year location matrices for every individual from their year of birth through 2023. When participants reported more than one residence in a given year, two variables were created:

- Former Location: Represents where the individual resided earlier or concurrently during that year.
- Latter Location: Reflects any subsequent or overlapping location reported for that same year.

This approach preserved temporal information and minimized the loss of data in cases of reporting more than one location.

### Grouping Countries into Locations

For analytical purposes, all locations were then classified into one of three categories:

- American Samoa
- Tropical Islands (e.g., Samoa, Tuvalu, Fiji, Philippines)
- Non-endemic areas history (e.g., Continental US, Hawaii, New Zealand and Australia)

These categories enabled epidemiological comparisons across endemic and non-endemic exposure environments. The finalized longitudinal dataset provided a structured framework for evaluating residence-based risk in the serosurvey cohort.

Most results shown in the paper regarding residency data are using the dataset where the “former” locations are considered the correct ones.

### Catalytic model to estimate force of infection and catalytic model regression analysis

We assumed that an individual’s seropositivity follows a Bernoulli distribution, with the probability of seropositivity determined by the cumulative hazard of DENV infection over their lifetime. Specifically, we defined the annual hazard of DENV infection, averaged across the four serotypes, for an individual residing in American Samoa in year *t* as 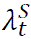, and for individuals residing in dengue non-endemic or endemic countries as 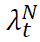 and 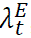, respectively. At the time of the serosurvey (*t=Y*), an individual *i* of age *a* and residence history 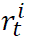 experienced cumulative dengue FOI, calculated as: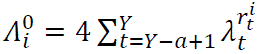. We assumed that hazard rates for non-endemic and endemic countries were constant over time and across countries. Finally, we assumed that dengue hazard varied by school according to a multiplicative random effect following a Gamma distribution with mean one and variance 1/α to be estimated.

To account for the complex survey design, we assumed that all individuals were independent and that the sample was representative with respect to characteristics not already included in the survey weights. Model fitting was performed using a likelihood function weighted by the final design weights. Specifically, the log-likelihood contribution of participant *i* of age *a* with residence history 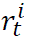 attending school *s*, with serostatus *y_i_*=1 for seropositive individuals and *y*_1_ = 0 for seronegative individuals, is 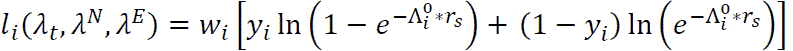, with 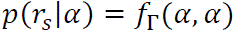, where *w*_i_ is the survey weight.

To account for imperfect sensitivity and specificity of the diagnostic test, we modeled observed seropositivity as a function of both true positives (seropositive individuals who test positive) and true negatives (seronegative individuals who test negative). Sensitivity and specificity estimates were inferred from the number of positive and negative tests among well characterized positive and negative samples from an internal validation study at CDC Dengue Branch.(22) The log-likelihood of the validation study was included using a binomial distribution: given TP, FN, FP, and TN true positives, false negatives, false positives, and true negatives respectively, *l*_*validation*_(*se*, *sp*) = *TP* ln(*se*) + *FN* ln(1 − *se*) + *FP* ln(1 − *sp*) + *TN* ln(*sp*).

We incorporated an independent serosurvey conducted in American Samoa among adults aged 18-40 in 2010 to better inform historical seroprevalence. The model-estimated, age-stratified seroprevalence was constructed using historical FOI (providing estimates π_18*to*25_ and π_26*to*40_ for the age groups 18 to 25 and 26 to 40), and a binomial distribution was used to model the likelihood of observing the seroprevalence in the study, given the model-estimated seroprevalence. Given a total of p_18to25_ seropositive individuals out of N_18to25_ and p_26to40_ seropositive individuals out of N_26to40_ surveyed in 2010 in age groups 18-25 and 26-40 respectively, the likelihood contribution of the 2010 serosurvey was *l*_2010_(**λ**) = *p*_18*to*25_ ln(π_18*to*25_) + (*N*_18*to*25_ − *p*_18*to*25_) ln(1 − π_18*to*25_) + *p*_26*to*40_ ln(π_26*to*40_) + (*N*_26*to*40_ − *p*_26*to*40_) ln(1 − π_26*to*40_).

Finally, we combined individual-level seropositivity data with age-stratified dengue case data from American Samoa to jointly estimate FOI. Following the approach of Kada et al. (25), we derived the likelihood for the annual number of RT-PCR-confirmed dengue cases reported in each age group from 2016 to 2023 using ArboNET data. For the years covered by the serosurvey, we estimated the FOI in as a survey-weighted mean FOI among serosurvey participants to account for sample non-representativeness. The FOI was then used to calculate the expected number of cases in each age group, assuming that cases represent a mixture of primary and secondary infections. We estimated annual reporting rate, and the relative reporting rate for primary vs. secondary infections. We jointly maximized the sum of the weighted log-likelihood from the serosurvey, the log-likelihood from the 2010 serosurvey, and the log-likelihood from the age-stratified case data. Parameter estimation was performed using Markov chain Monte Carlo (MCMC) methods implemented in the rstan package. We report median parameter estimates and associated 95% credible intervals from the posterior distributions.

## Notes

### Competing Interest Statement

The authors have declared no competing interest.

### Funding Statement

This study did not receive any external funding.

### Author Declarations

The Centers for Disease Control and Prevention determined this work to be non-human subjects research as it met criteria for public health surveillance exclusion. The datasets used in the study were either individual-level data that was de-identified or in an aggregated/summary data from a research paper.

